# Large-scale genome-wide interaction analyses on multiple cardiometabolic risk factors to identify age-specific genetic risk factors

**DOI:** 10.1101/2024.07.12.24310321

**Authors:** Linjun Ao, Diana van Heemst, Jiao Luo, Maris Teder-Laving, Reedik Mägi, Estonian Biobank Research Team, Ruth Frikke-Schmidt, Ko Willems van Dijk, Raymond Noordam

**Affiliations:** Department of Human Genetics, Leiden University Medical Center, Leiden, the Netherlands; Department of Internal Medicine, Section of Gerontology and Geriatrics, Leiden, the Netherlands; Department of Clinical Biochemistry, Copenhagen University Hospital-Rigshospitalet, Copenhagen, Denmark; Estonian Genome Center, Institute of Genomics, University of Tartu, Tartu, Estonia; The Copenhagen General Population Study, Herlev and Gentofte Hospital, Herlev, Denmark; Department of Clinical Medicine, Faculty of Health and Medical Sciences, University of Copenhagen, Copenhagen, Denmark; Department of Internal Medicine, Division of Endocrinology, Leiden University Medical Center, Leiden, the Netherlands; Einthoven Laboratory for Experimental Vascular Medicine, Leiden University Medical Center, Leiden, the Netherlands

## Abstract

**Background:** The genetic landscape of cardiometabolic risk factors has been explored extensively. However, insight in the effects of genetic variation on these risk factors over the life course is sparse. Here, we performed genome-wide interaction studies (GWIS) on different cardiometabolic risk factors to identify age-specific genetic risks.

**Methods:** This study included 270,276 unrelated European-ancestry participants from the UK Biobank (54.2% women, a median age of 58 [interquartile range (IQR): 50, 63] years). GWIS models with interaction terms between genetic variants and age were performed on apolipoprotein B (ApoB), low-density lipoprotein-cholesterol (LDL-C), log-transformed triglycerides (TG), body mass index (BMI), and systolic blood pressure (SBP). Replication was subsequently performed in the Copenhagen General Population Study (CGPS) and the Estonian Biobank (EstBB).

**Results:** Multiple lead variants were identified to have genome-wide significant interactions with age (*P_interaction_* <1e-08). In detail, rs429358 (tagging *APOE4*) was identified for ApoB (*P_interaction_* = 9.0e-14) and TG (*P_interaction_* = 5.4e-16). Three additional lead variants were identified for ApoB: rs11591147 (R46L in *PCSK9*, *P_interaction_*= 3.9e-09), rs34601365 (near *APOB*, *P_interaction_* = 8.4e-09), and rs17248720 (near *LDLR*, *P_interaction_* = 2.0e-09). Effect sizes of the identified lead variants were generally closer to the null with increasing age. No variant-age interactions were identified for LDL-C, SBP and BMI. The significant interactions of rs429358 with age on ApoB and TG were replicated in both CGPS and EstBB.

**Conclusions:** The majority of genetic effects on cardiometabolic risk factors remains relatively constant over age, with the noted exceptions of specific genetic effects on ApoB and TG.

## Introduction

Cardiovascular disease (CVD) remains a leading cause of death worldwide, and contributes substantially to morbidity and healthcare costs (1, 2). It is widely recognized that dyslipidaemia, hypertension, obesity, and behavioural factors such as smoking are important cardiovascular risk factors (2, 3). With the expansion of human genetic datasets, genome-wide association studies (GWAS) have provided increasing insight into the underlying biological mechanisms of risk factors for multifactorial diseases which has resulted in the identification of targets for cardiovascular risk management and CVD prevention (4–6). Also, the Global Lipids Genetics Consortium (GLGC) identified several novel and ancestry-specific loci for dyslipidaemia, resulting in improved insight in the underlying biology and fine-mapping of functional variants (7, 8).

Most cardiometabolic risk factors are influenced by a combination of genetic and non-genetic factors (9–11). Age is an important non-modifiable determinant for CVD risk (12, 13). Several studies have reported that the relative impact of modifiable risk factors on CVD risk may be greater in younger than in older individuals (14–16). However, the impact of age on the genetic architecture of cardiovascular risk factors has not been widely explored yet (12), which may be an explanation of the attenuated associations with increasing age. As the number of people reaching advanced age is increasing, the investigation of interactions between genetic variation and age on cardiovascular risk factors is increasingly important for the identification of targets for CVD prevention and intervention in older people.

Cardiometabolic risk factors, including dyslipidaemia, hypertension, and obesity are predominant risk factors for CVD (17–19). Few studies have examined the interactions of genetic variants with age on blood pressure and body mass index (BMI), and only a few variants with small effect sizes varying over the life time have been identified thus far (20, 21). Increased low-density lipoprotein-cholesterol (LDL-C) and triglycerides (TG) are main components of dyslipidaemia associated with CVD risk (19, 22, 23). Recently, apoprotein B (ApoB) has been identified as a more precise indicator of CVD risk than LDL-C (24, 25). Thus far, insight in the effects of genetic variation on cardiometabolic risk factors over the life course is limited. Therefore, we aimed to assess the interactions of genetic variants with age on common cardiometabolic risk factors, namely ApoB, LDL-C, TG, BMI, and systolic blood pressure (SBP), by large-scale genome-wide interaction analysis (GWIS).

## Methods

### Study population and design

The primary (discovery) analyses of the present study were embedded in the prospective UK Biobank (UKB) cohort, which recruited over 500,000 participants aged 40-70 years across the entire United Kingdom during the baseline survey between 2006 and 2010. Extensive phenotypic and genotypic details of the participants have been collected since the baseline assessment, including sociodemographic data, lifestyle, physical measures, biological samples (blood, urine and saliva), genome-wide genotyping, and longitudinal follow-up on a wide range of health-related outcomes. The UKB cohort study was approved by the North-West Multicentre Research Ethics Committee (MREC). All participants provided electronic written informed consent for the study. A detailed description of the UKB cohort study has been presented elsewhere (26).

To minimize population stratification bias, the present study restricted participants to 318,734 unrelated individuals with European ancestry, based on the estimated kinship coefficients for all pairs and the self-reported ancestral background (27). After excluding individuals with missing data on the examined five risk factors, we ultimately included 270,276 participants. Details of missingness for each trait are presented in supplementary Table S1, with the largest percentage of missingness for SBP being 8.7%.

### Cardiometabolic risk factors

All five cardiometabolic risk factors, being ApoB (g/L), LDL-C (mmol/L), TG (mmol/L), BMI (kg/m^2^), and SBP (mmHg), were collected and measured during the baseline assessment. ApoB, LDL-C, and TG were measured based on blood samples with the Beckman Coulter AU5800. Consistent with studies conducted by some large consortia (8, 28), the LDL-C level was divided by 0.7 if participants used statins. TG was natural log-transformed to normal distribution for subsequent analyses. The BMI values in the UKB data were calculated from height and weight. SBP was measured twice in a resting sitting position at the study centre, and the average of the two measurements was used. In agreement with previous studies, including genetic studies (21), if participants reported taking antihypertensive medication, 10 mmHg were added to the mean of the measured SBP. Besides, if a value was more than 6 standard deviations (SD) above or below the mean, we set it to exactly at 6 SDs from the mean.

### Genotyping and genetic imputations

UKB genotyping was conducted by Affymetrix using a bespoke BiLEVE Axium array for approximately 50,000 participants, and using the Affymetrix UK Biobank Axiom array for the remaining participants. All genetic data were quality controlled centrally by UKB resources. More information on the genotyping processes can be found online (https://www.ukbiobank.ac.uk). Based on the genotyped single-nucleotide polymorphisms (SNPs), UKB resources performed centralized imputations on the autosomal SNPs using the UK10K haplotype (29), 1000 Genomes Phase 3 (30), and Haplotype Reference Consortium reference panels (31). Autosomal SNPs were pre-phased using SHAPEIT3 and imputed using IMPUTE4. In total, ∼96 million SNPs were imputed.

### Genome-wide interaction analyses

Using the software program GEM (version 1.4.2) (32), the GWIS of each cardiovascular risk factor was carried out for the included 270,276 UKB individuals by the generalized linear model, with covariates including age, sex, first ten genetic principal components (PCs), and an interaction term between genetic variant and age. SNPs with a minor allele frequency below 0.001 were removed. The genome-wide significant interaction effect was set at a *P* value less than 1e-8 (5e-8 /five risk factors) to correct for the multiple testing. We used the Functional Mapping and Annotation of Genome-Wide Association Studies (FUMA) web-based application (https://fuma.ctglab.nl/) (33) to identify independent lead genetic variants (r^2^ < 0.1), using the 1000 G Phase 3 EUR as reference panel population. Positional mapping is performed based on annotations obtained from ANNOVAR (34) with the maximum distance of 10kb from genetic variants to genes.

### Look-up analyses for potential gene-age interactions

The power issues for strict genome-wide significant tests may result in some variants with weak interactions with age not being identified. In addition to the primary analyses, we thus performed conventional (marginal) GWAS, and extracted independent lead variants with genome-wide significant effects on the corresponding risk factor (*P*-values for the marginal effects less than 5e-8). Subsequently, we explored their interaction effects with age in the GWIS described above. The statistically significant threshold of the interaction term was defined as 0.05 divided by the corresponding number of extracted genetic variants for each risk factor. The identified variants showing statistically significant interaction with age were then explored in GWAS Catalogue (https://www.ebi.ac.uk/gwas/) to investigate their mapped gene.

### Stratified analyses for lead genetic variants

Included participants were categorized into three age groups, [40, 50), [50, 60) and [60,70] years. For lead genetic variants showing genome-wide significant interaction with age after Bonferroni correction (*P*-values for the interaction terms less than 1e-8), we performed linear regressions to assess the associations of their genotypes with the corresponding risk factors in the different age groups, adjusting for sex and the first ten genetic PCs. We further tested the interaction effects as well as the age-stratified effects of the identified lead variants and age in women and men separately.

### Replication of the main study results

The Copenhagen General Population Study (CGPS) is an ongoing prospective cohort study of 109,751 Danish adults aged 20–100 years, recruited between 2003 and 2015 (35). Invited individuals were randomly selected from the national Danish Civil Registration System to represent the general population of white Danish adults. All participants filled in a questionnaire, had a physical examination, and had blood samples collected for biochemical analyses at the baseline survey.

The Estonian Biobank (EstBB) is a population-based biobank cohort that currently comprises more than 200,□000 individuals, representing ∼ 20% of the adult population in Estonia. Details of the EstBB has been described elsewhere (36). Briefly, all included participants completed a comprehensive questionnaire at baseline, including personal data, genealogical data, lifestyle data, medical history and current health status, etc. Blood samples for DNA, plasma, and white blood cell are also collected and stored at baseline. Besides, all EstBB participants have been genotyped. The EstBB project is being conducted according to the Estonian Human Genes Research Act (HGRA), and all included participants have signed a broad informed consent form.

For replication purposes of the main findings, in CGPS and EstBB, we tested the interactions between the lead variants and age, and performed the age-stratified analyses. For analyses conducted in EstBB, generalised linear models were adjusted for sex, age (not included in age-stratified analyses) and the first ten genetic PCs. As only a small proportion of participants have chip data in CGPS, the generalised linear models in CGPS were unadjusted for genetic PCs and only adjusted for sex and age. In EstBB and CGPS, interactions were also tested separately for women and men, and was carried out in the sub-population of 40-to 70-year-olds to align with the UKB study population.

## Results

### Characteristics of study participants

A total of 270,276 unrelated European-ancestry participants (54.2% women, and a median age at inclusion of 58 [interquartile range (IQR): 50, 63] years) from UKB were eligible for analyses in this study. The baseline characteristics of the cardiometabolic risk factors in the UKB stratified by age, are presented in Table 1. In addition, 97,283 participants (65.6% women) from EstBB, and 107,435 participants (55.11% women) for ApoB and 107,504 participants (55.10% women) for TG from CGPS were included for validation analyses. The detailed characterises from both two validation studies were presented in Table S2. In general, and as expected, the levels of the examined risk factors were higher in the older group.

**Table 1.** The baseline characteristics of the study population from UK Biobank.

|  | **Overall** | **[40, 50)** | **[50, 60)** | **[60, 70]** |
| --- | --- | --- | --- | --- |
| n | 270276 | 67989 | 95695 | 106592 |
| Age (median [IQR]) | 58 [50, 63] | 46 [43, 48] | 56 [53, 59] | 64 [62, 67] |
| Sex = Male, n (%) | 123805 (45.8) | 30901 (45.4) | 42116 (44.0) | 50788 (47.6) |
| ApoB (g/L), mean (SD) | 1.03 (0.24) | 1.00 (0.23) | 1.06 (0.23) | 1.04 (0.24) |
| LDL-C (mmol/L), mean (SD) | 3.57 (0.86) | 3.46 (0.79) | 3.66 (0.84) | 3.56 (0.90) |
| TG (mmol/L), median [IQR] | 1.49 [1.05, 2.14] | 1.31 [0.92, 2.00] | 1.50 [1.06, 2.17] | 1.57 [1.14, 2.20] |
| SBP (mmHg), mean (SD) | 137.86 (18.57) | 129.54 (15.85) | 136.95 (17.76) | 143.98 (18.66) |
| BMI (kg/m<sup>2</sup>), mean (SD) | 27.34 (4.70) | 26.92 (4.85) | 27.43 (4.84) | 27.53 (4.46) |
| Lipid-lowering medication = 1, n (%) | 45929 (17.0) | 2844 ( 4.2) | 13085 (13.7) | 30000 (28.1) |
| BP-lowering medication = 1, n (%) | 54693 (20.2) | 4133 ( 6.1) | 16822 (17.6) | 33738 (31.7) |

Abbreviations: ApoB, apolipoprotein B; BMI, body mass index; BP, blood pressure; IQR: interquartile range; LDL-C, low-density lipoprotein cholesterol; SBP, systolic blood pressure; SD, standard deviation; TG, triglyceride.

In addition, the genotype frequencies of the lead genetic variants (detailed below) in different age groups from the UKB and validation studies (EstBB and CGPS) were presented in Table 2. The frequencies of the examined genotypes were similar across the different age groups.

**Table 2.** The genotype distribution of the identified lead variants among different age groups in UK Biobank and Validation cohorts.

|  |  | UK Biobank |  |  | Estonian Biobank |  |  |  |  | Copenhagen General Population Study |  |  |  |  |  |
| --- | --- | --- | --- | --- | --- | --- | --- | --- | --- | --- | --- | --- | --- | --- | --- |
|  |  | [40, 50) | [50, 60) | [60, 70] | [20, 40) | [40, 50) | [50, 60) | [60, 70) | [70, 80] | [20, 40) | [40, 50) | [50, 60) | [60, 70) | [70, 80) | [80, 110] |
| rs11591147 | G; G | 96.62% | 96.56% | 96.5% | 97.06% | 97.19% | 97.04% | 96.83% | 96.93% | 97.77% | 97.35% | 97.34% | 97.32% | 97.32% | 96.99% |
|  | T; G | 3.35% | 3.41% | 3.47% | 2.91% | 2.76% | 2.93% | 3.14% | 3.03% | 2.21% | 2.63% | 2.64% | 2.66% | 2.67% | 3.01% |
|  | T; T | 0.03% | 0.03% | 0.03% | 0.03% | 0.05% | 0.03% | 0.03% | 0.05% | 0.02% | 0.02% | 0.02% | 0.02% | 0.01% | 0.00% |
| rs17248720 | C; C | 77.74% | 77.7% | 77.94% | 84.33% | 84.40% | 84.54% | 84.28% | 84.52% |  |  |  |  |  |  |
|  | T; C | 20.91% | 20.94% | 20.65% | 15.00% | 14.92% | 14.82% | 15.00% | 14.87% |  |  |  |  |  |  |
|  | T; T | 1.35% | 1.36% | 1.41% | 0.68% | 0.67% | 0.64% | 0.73% | 0.61% |  |  |  |  |  |  |
| rs34601365* | C; C | 4.08% | 4.06% | 4.04% | 11.14% | 11.32% | 11.17% | 11.35% | 10.60% |  |  |  |  |  |  |
|  | C; CT | 32.06% | 32.24% | 32.42% | 44.19% | 44.63% | 44.29% | 43.84% | 44.91% |  |  |  |  |  |  |
|  | CT; CT | 63.86% | 63.7% | 63.54% | 44.67% | 44.06% | 44.54% | 44.81% | 44.49% |  |  |  |  |  |  |
| rs429358 | T; T | 71.08% | 71.48% | 71.98% | 76.30% | 75.87% | 75.88% | 76.22% | 75.94% | 67.58% | 68.28% | 68.27% | 68.26% | 70.36% | 72.63% |
|  | C; T | 26.52% | 26.17% | 25.71% | 22.01% | 22.53% | 22.40% | 21.92% | 22.75% | 29.15% | 28.61% | 28.70% | 28.83% | 26.97% | 25.72% |
|  | C; C | 2.40% | 2.34% | 2.30% | 1.69% | 1.60% | 1.72% | 1.87% | 1.30% | 3.28% | 3.11% | 3.03% | 2.91% | 2.67% | 1.65% |

\*: For Estonian Biobank, the table showed the genotype frequency of rs62122481(AA; AC; CC in descending order) as rs62122481 is an proxy of rs34601365 and mapped to the same *ApoB* gene.

### Genome-wide interaction analyses

In total, we observed genome-wide significant interaction effects (*P*-values for interaction terms < 5e-8) between 258 genetic variants and age on the examined phenotypes, of which 234 for ApoB, 23 for TG, and 1 for BMI (Table S3). No genome-wide significant gene-age interaction effects were identified for LDL-C and SBP. After Bonferroni correction for multiple testing for the number of examined phenotypes, 70 variants remained that had genome-wide significant interaction effects with age (*P*-values for interaction terms < 1e-8), of which 48 for ApoB and 22 for TG.

Among these 70 variants with significant interactions with age, four lead variants for ApoB (rs11591147 (*P_interaction_* = 3.9e-09, β_interaction_ = 0.0018) mapping to *PCSK9*; rs34601365 (*P_interaction_* = 8.4e-09, β_interaction_ = -0.0006) mapping to *TDRD15*; rs17248720 (*P_interaction_* = 2.0e-09, β_interaction_ = 0.0007) mapping to *LDLR*; and rs429358 (*P_interaction_* = 9.0e-14, β_interaction_ = - 0.0009) mapping to *PVRL2*, *TOMM40*, *APOE*, and *APOC1*), and one lead variant for TG (rs429358 (*P_interaction_*= 5.4e-16, β_interaction_ = -0.0019) mapping to *PVRL2*, *TOMM40*, *APOE*, and *APOC1*) were identified (Figure 1).

**Figure 1.**
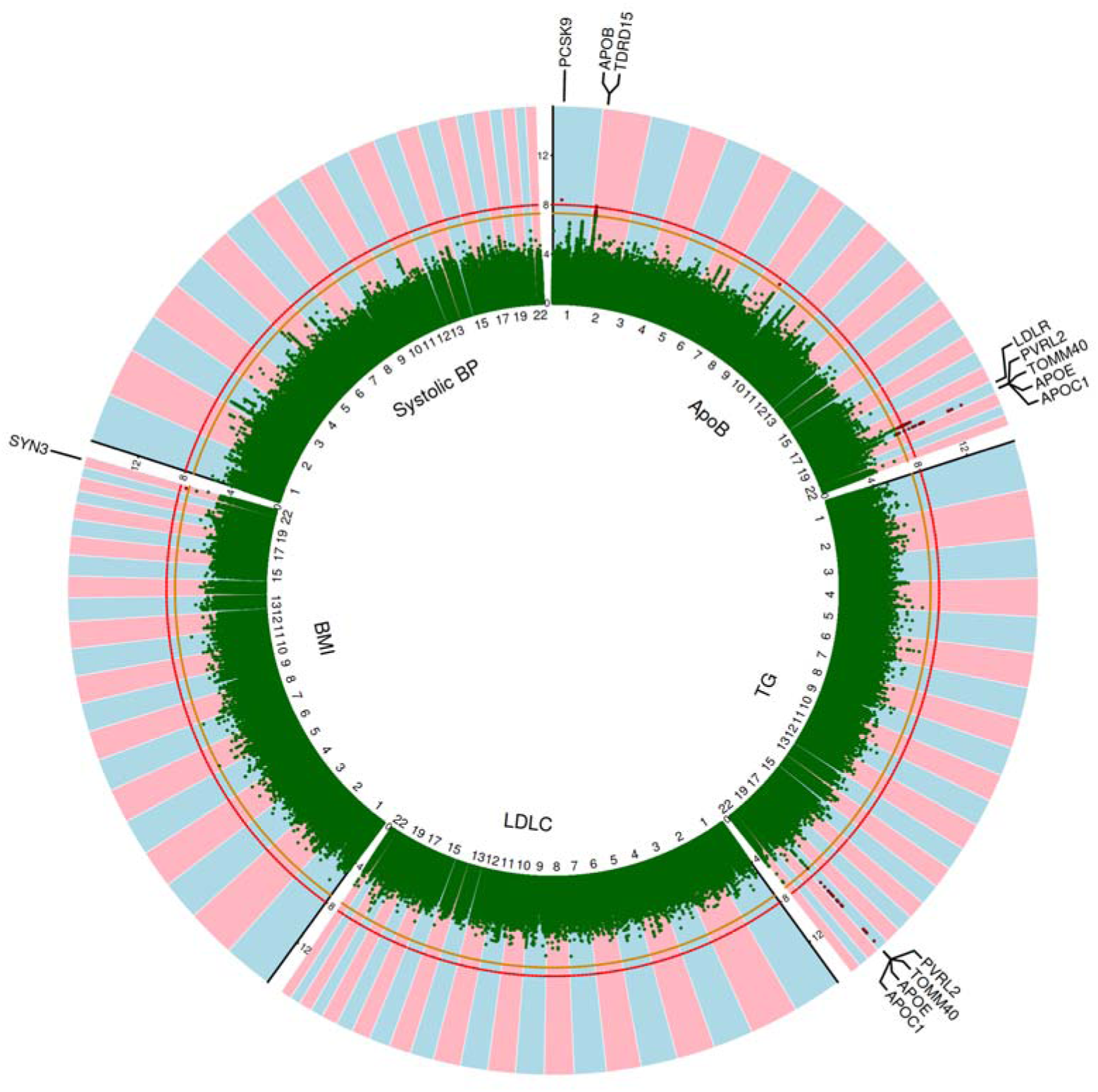
Circular Manhattan plot for the log-transformed (base 10) *P*-values of the interaction terms between genetic variants and age. Abbreviations in clockwise direction: ApoB, apolipoprotein B; TG, triglyceride; LDLC, low-density lipoprotein cholesterol; BMI, body mass index; BP, blood pressure. The orange line indicates a *P*-value of 5e-8, and the red line indicates a *P*-value of 1e-8 after Bonferroni correction for multiple testing. Red dots indicate genome-wide significant SNPs with *P*-values smaller than 5e-8 for the interaction terms. Labelled gene names in black were identified by FUMA.

Except for the interaction of rs17248720 with age on ApoB in women, the interactions of the lead variants with age remained significant (*P*-values for the interaction terms < 0.01) in both women and men (Table S4). In addition, the interaction results from validation cohorts (EstBB and CGPS) were presented in Table S5. Notably, both two cohorts showed significant interactions between rs429358 [tagging *APOE4*] and age on ApoB (*P_interaction_* = 4.60e-05 in EstBB; *P_interaction_* = 9.07e-05 in CGPS) and on TG (*P_interaction_* = 2.59e-05 in EstBB; *P_interaction_* =2.35e-07 in CGPS). These interactions remained in both women and men, and in the 40-70 year old subpopulation. (Table S5).

### Look-up analyses for potential gene-age interactions

A total of 958 independent genetic variants showed marginal effects on the corresponding risk factor, of which 145 were associated with ApoB, 175 with TG, 198 with LDL-C, 239 with BMI and 201 with SBP. Among these genetic variants, a total of 17 independent variants showed evidence for interaction with age after correction for multiple testing, i.e., 9 for ApoB (*P*-values for the interaction terms < 0.05/145), 2 for TG (*P*-values for the interaction terms < 0.05/175), 1 for LDL-C (*P*-values for the interaction terms < 0.05/198), 3 for BMI (*P*-values for the interaction terms < 0.05/239), and 2 for SBP (*P*-values for the interaction terms < 0.05/201). In addition to already identified genes by GWIS, several more genes were found, such as *LIPC* for ApoB (rs261334, *P*_interaction_ = 4.44e-06) and TG (rs1077835, *P*_interaction_ = 1.16e-04), and *FTO* (rs11642015, *P*_interaction_ = 1.1e-04) for BMI (Table 3).

**Table 3.**
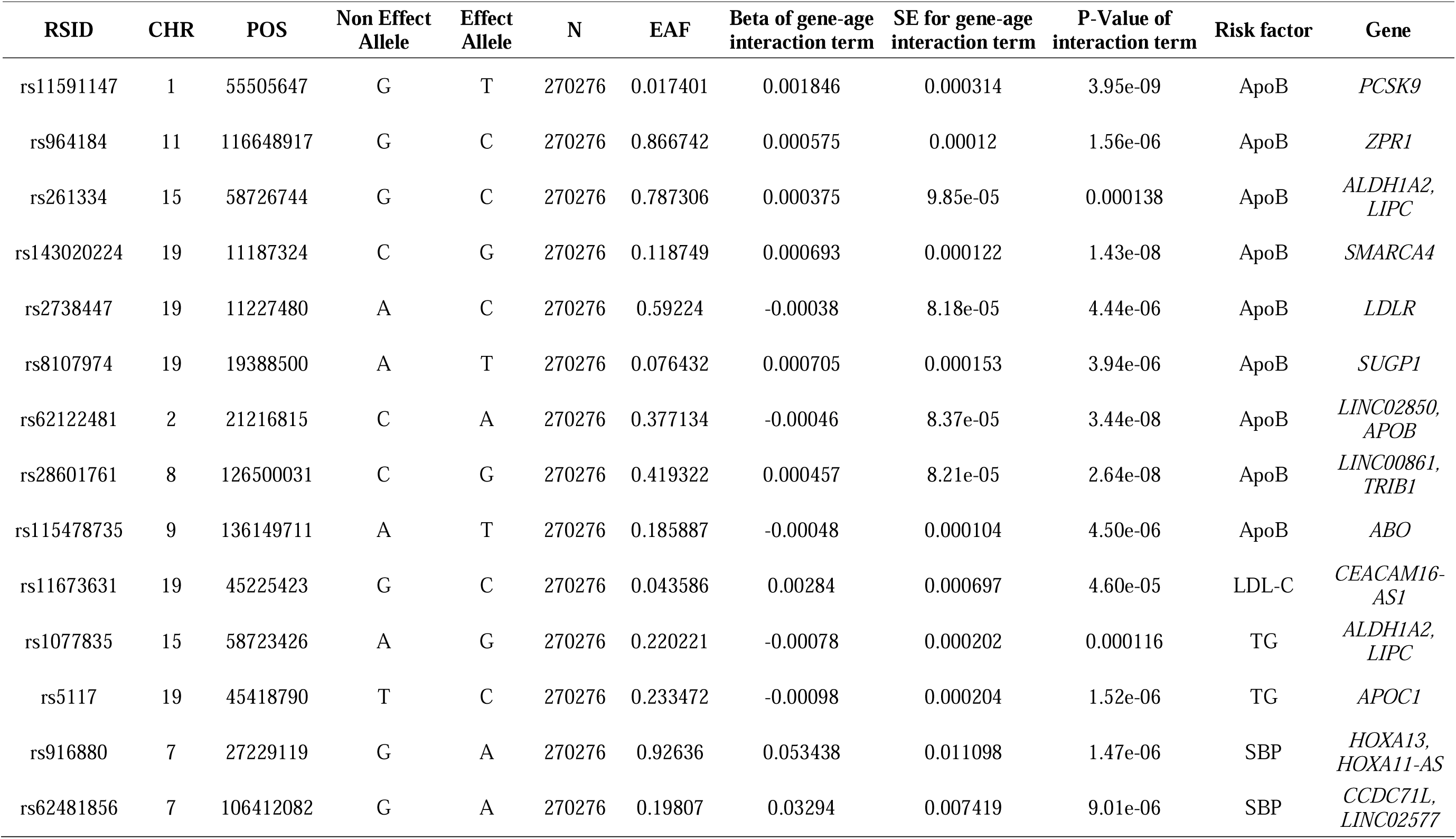
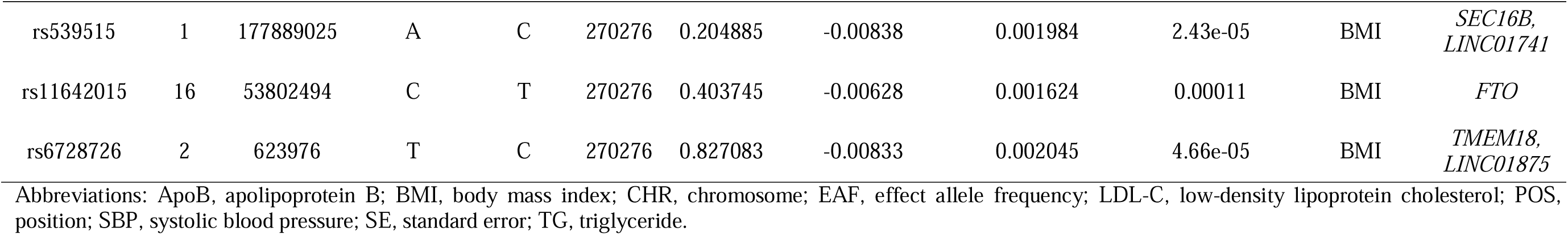
Genetic variants showing statistically significant interaction with age by look-up analyses.

| RSID | CHR | POS | Non Effect Allele | Effect Allele | N | EAf | Beta of gene-age interaction term | SE for gene-age interaction term | P-Value of interaction term | Risk factor | Gene |
| --- | --- | --- | --- | --- | --- | --- | --- | --- | --- | --- | --- |
| rs11591147 | 1 | 55505647 | G | T | 270276 | 0.017401 | 0.001846 | 0.000314 | 3.95e-09 | ApoB | *PCSK9* |
| rs964184 | 11 | 116648917 | G | C | 270276 | 0.866742 | 0.000575 | 0.00012 | 1.56e-06 | ApoB | *ZPR1* |
| rs261334 | 15 | 58726744 | G | C | 270276 | 0.787306 | 0.000375 | 9.85e-05 | 0.000138 | ApoB | *ALDH1A2, LIPC* |
| rs143020224 | 19 | 11187324 | C | G | 270276 | 0.118749 | 0.000693 | 0.000122 | 1.43e-08 | ApoB | *SMARCA4* |
| rs2738447 | 19 | 11227480 | A | C | 270276 | 0.59224 | -0.00038 | 8.18e-05 | 4.44e-06 | ApoB | *LDLR* |
| rs8107974 | 19 | 19388500 | A | T | 270276 | 0.076432 | 0.000705 | 0.000153 | 3.94e-06 | ApoB | *SUGP1* |
| rs62122481 | 2 | 21216815 | C | A | 270276 | 0.377134 | -0.00046 | 8.37e-05 | 3.44e-08 | ApoB | *LINC02850, APOB* |
| rs28601761 | 8 | 126500031 | C | G | 270276 | 0.419322 | 0.000457 | 8.21e-05 | 2.64e-08 | ApoB | *LINC00861, TRIB1* |
| rs115478735 | 9 | 136149711 | A | T | 270276 | 0.185887 | -0.00048 | 0.000104 | 4.50e-06 | ApoB | *ABO* |
| rs11673631 | 19 | 45225423 | G | C | 270276 | 0.043586 | 0.00284 | 0.000697 | 4.60e-05 | LDL-C | *CEACAM16-AS1* |
| rs1077835 | 15 | 58723426 | A | G | 270276 | 0.220221 | -0.00078 | 0.000202 | 0.000116 | TG | *ALDH1A2, LIPC* |
| rs5117 | 19 | 45418790 | T | C | 270276 | 0.233472 | -0.00098 | 0.000204 | 1.52e-06 | TG | *APOC1* |
| rs916880 | 7 | 27229119 | G | A | 270276 | 0.92636 | 0.053438 | 0.011098 | 1.47e-06 | SBP | *HOXA13, HOXA11-AS* |
| rs62481856 | 7 | 106412082 | G | A | 270276 | 0.19807 | 0.03294 | 0.007419 | 9.01e-06 | SBP | *CCDC71L, LINC02577* |
| rs539515 | 1 | 177889025 | A | C | 270276 | 0.204885 | -0.00838 | 0.001984 | 2.43e-05 | BMI | *SEC16B,*<br>*LINC01741* |
| rs11642015 | 16 | 53802494 | C | T | 270276 | 0.403745 | -0.00628 | 0.001624 | 0.00011 | BMI | *FTO* |
| rs6728726 | 2 | 623976 | T | C | 270276 | 0.827083 | -0.00833 | 0.002045 | 4.66e-05 | BMI | *TMEM18,*<br>*LINC01875* |

Abbreviations: ApoB, apolipoprotein B; BMI, body mass index; CHR, chromosome; EAF, effect allele frequency; LDL-C, low-density lipoprotein cholesterol; POS, position; SBP, systolic blood pressure; SE, standard error; TG, triglyceride.

### Stratified analyses

Figure 2 shows the associations of the four lead SNPs (rs11591147, rs34601365, rs17248720, rs429358) with ApoB and the association of rs429358 with TG in different age groups. The homozygous genotypes were observed to have greater effects on the corresponding risk factors than the heterozygous genotypes. Notably, with the exception of the association between the homozygous group of rs11591147 [R46L in *PCSK9*] and ApoB, the associations of the genotype groups (both heterozygous and homozygous) relative to the reference group with the corresponding phenotypes attenuated with age. For example, the homozygous genotype (C; C) of rs429358 [tagging *APOE4*] had the largest effect on TG in the 40-to 50-year-old age group, with a 1.11-fold [95% CI: 1.08, 1.14] increase, and had the smallest effect in the 60-to 70-year-old age group, with a 1.03-fold [95% CI: 1.01, 1.05] increase.

**Figure 2.**
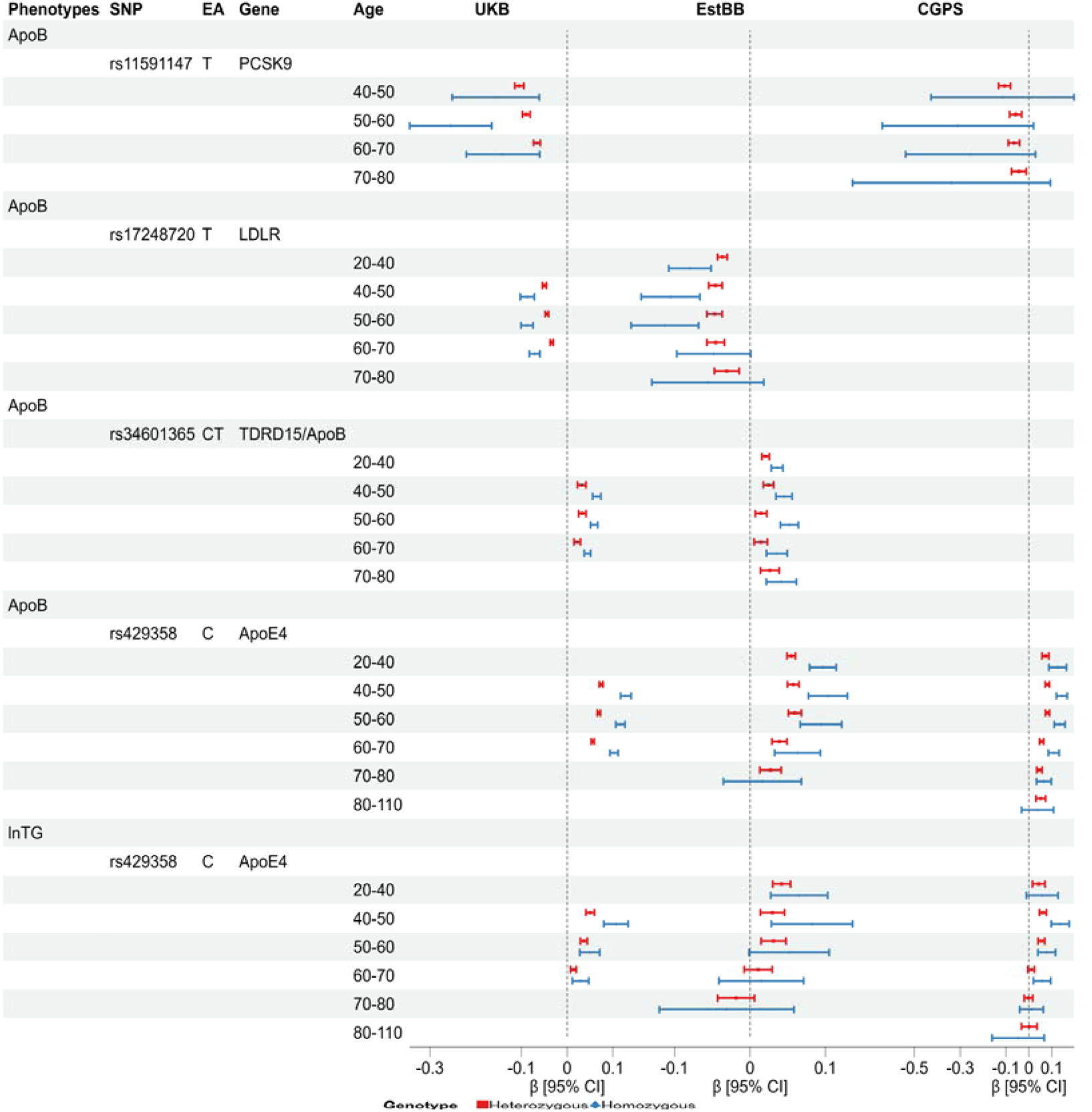
Associations between the genotypes of the lead SNPs and the corresponding phenotypes for age-stratified analyses in UKB and two validation cohorts. ApoB, apolipoprotein B; EA, effect allele; SNP: single nucleotide polymorphism; TG: triglyceride. Cohort names: UKB, UK Biobank; EstBB, Estonian Biobank; CGPS, Copenhagen General Population Study. In UKB and EstBB, linear regressions were adjusted for sex and the first ten genetic principal components, whereas in CGPS only sex was adjusted. For EstBB, the figure showed the results of rs62122481 (effect allele: A), which is an proxy SNP for rs34601365.

The results of the age-stratified analyses for women and men from UKB were similar to those of the main analysis (Figure S1). In addition, the age-stratified analyses in validation cohorts showed similar results to the main analyses (Figure 2). The direction and the decreasing trend with aging for all the genetic effects, especially for rs429358, are in line with the main analyses.

## Discussion

Our genome-wide interaction studies in 270,276 unrelated European-ancestry participants from UKB identified multiple genetic variants that showed significant interactions with age on two of the five examined cardiometabolic risk factors. Specifically, four lead variants were identified for ApoB: rs11591147 [R46L in *PCSK9*], rs34601365 [near *TDRD15* and *APOB*], rs17248720 [near *LDLR*], and rs429358 [tagging *APOE4*]; one lead variant was identified for TG: rs429358 [tagging *APOE4*]. No genome-wide significant interaction effects were found for LDL-C, SBP and BMI. The effect sizes of identified lead variants were closer to null with increasing age. The interactions of rs429358 [tagging *APOE4*] with age were replicated in EstBB and CGPS.

In the present study, three independent variants, which are located near or in genes that play a clear role in ApoB metabolism, were found to have significant interactions with age on ApoB levels. First, it is well known that the proprotein convertase kexin/subtilisin type 9 (PCSK9) increases plasma levels of LDL-C by interacting with the LDL receptor (LDLR) and decreasing endocytic recycling of the LDLR. The missense R46L variant in the *PCSK9* gene is thought to inhibit this cycle and thereby lower LDL-C (37, 38). A previous study showed that carriers of *PCSK9* R46L variant could lower LDL-C level and ApoB level (39). In addition, the common variant rs17248720-T, located at the 5’ end of *LDLR* gene, was found to be associated with increased *LDLR* transcriptional activity, lower LDL-C levels (40), and lower non-high-density lipoproteins (non-HDL) cholesterol levels (41). These observations are in line with changes in ApoB levels. In accordance, we found that both the R46L variant in *PCSK9* and the rs17248720-T variant near the *LDLR* gene were associated with the lower ApoB levels. Since ApoB-100 is the main structural protein of LDL, the lower LDL-C levels caused by the R46L variant and rs17248720-T variant is therefore likely paralleled by reduced ApoB levels. Lastly, rs34601365 is in vicinity of the *TDRD15* and *APOB* genes, which are located in the same genomic locus, less than 250kb apart. There is evidence that the rs34601365 or its proxy SNP rs62122481 (effect allele: A, mapping to *APOB*) associated with higher ApoB levels (42).

Interestingly, our study, including the replication analyses, found that rs429058, tagging the *APOE4*, had significant interactions with age on both ApoB and TG levels. Apolipoprotein E (ApoE) is an apolipoprotein component of chylomicrons, very-low-density lipoproteins (VLDL) and HDL. ApoE plays an important role as ligand responsible for the clearance of chylomicron and VLDL remnants in the liver through interaction with hepatic lipoprotein receptors, primarily the LDLR (43, 44). Among the three ApoE isoforms (ε2, ε3, and ε4), encoded by different *APOE* alleles, ApoE ε3 is the most common isoform. Relative to ApoE ε3, the ApoE ε4 has been found to be associated with higher TG, ApoB, and LDL-C levels (45). This has been attributed to the preference of ApoE ε4 for VLDL, a higher ApoE ε4-associated VLDL-production rate and/or higher VLDL-TG-lipolysis activity (46). In addition, participants with the homozygous *APOE* ε*4* have a lower hepatic LDLR activity than individuals with homozygous *APOE* ε*3* (47), which could lead to reduced clearance of VLDL remnants and, consequently, to TG accumulation as well. Our findings of elevated levels of ApoB and TG associated with the *APOE* ε*4* variant (rs429358, effect allele: C) are consistent with these results and conclusions.

It is well-known that the *APOE* ε*4* is negatively associated with human longevity (48). As shown in Table 2, the frequency of the *APOE* ε*4* allele was somewhat lower in older individuals (>70 years), and the interaction between rs429358 and age may be partially attributable to the negative effect of *APOE* ε*4* on longevity. In addition, we found that the genetic effects of all the identified variants on the corresponded phenotypes decreased with increasing age. There is evidence that the increase of LDL-C with age is explained by a reduced capacity for its removal, which would be mediated via a reduced hepatic LDLR expression (49). This finding has been validated by some animal studies (50). In conjunction with the roles of all the identified genes in lipoprotein metabolism as described above, the reduction in *LDLR* expression could also explain the attenuated genetic effects (including *APOE* ε*4* allele) with aging in our study.

Previous studies identified different genetic variants showing age-dependent effects on lipids levels during life course (51–53). For example, one study identified an age-dependent association (*P_interaction_*= 0.024) between rs2429917 [*SGSM2*] and LDL-C (50), while another study did not find any significant variants for LDL-C after the adjustment for multiple testing (52). In addition, one study of blood pressure using meta-regression models with a joint 2 degree of freedom likelihood ratio test identified 20 independent genetic variants exhibiting significant interactions with age, but none of those variants passed the interaction term test with a threshold of *P* < 5e-8 (21). Our study did not find genetic effects on SBP that varied significantly (*P* < 5e-8) across age, and only further identified 2 SNPs showing significant variant-age interactions (*P* < 0.00025) by look-up analyses (Table 3), one of which mapped to the same gene [*CCDC71L*] as found in the previous study (21). For the genetic effects on BMI over age, the present study identified one SNP with a threshold of *P* < 5e-8 (Figure 1), and further identified three SNPs with a threshold of *P* < 0.00021 by lookup analyses (Table 3). Three of the mapped genes i.e., *TMEM18*, *FTO*, and *SEC16B*, were also found in previous studies with a nominal significant threshold (*P_interaction_*< 0.05) (20, 54). However, it is important to note that, based on the studies mentioned above, there is little evidence of significant changes in genetic effects throughout the life course, which is concordant with our findings. Considering the increased prevalence of cardiovascular disease with aging (55, 56), all these findings may imply that the relative importance of genetic effects versus environmental influences could decrease with aging.

Mendelian randomization (MR) has emerged as a valuable tool to investigate potential causal associations by using genetic variants as instruments against false inferences resulting from confounding and reverse causality (57, 58). One of the assumptions to use MR methods is that the relationship between the genetic variant and the exposure should stay constant over time. Thus, the present study provides evidence that most genetic variants likely fulfil this key condition during adulthood. However, with the development of drug-targeted MR studies focusing on specific genetic variants (59), such as the effects of *PCSK9* inhibitor on atherosclerotic risk, caution must be exercised when combining or comparing results across studies with different age distributions.

The present study was conducted in a large study sample with a relatively large statistical power to detect genetic variants showing genome-wide significant interactions with age. In addition, our main findings were replicated in two other large cohort studies with a much larger age range. However, there are some limitations to be addressed. First, due to the lack of data for the number, dose, and type of antihypertensive medications taken, we could not correct the blood pressure parameters accurately. In addition, we screened for age-dependent genetic effects by incorporating an interaction term between variants and age in statistical models. This approach does not address the molecular mechanisms underlying the interactions in determining a phenotype, thereby potentially limiting insights into the biology.

In conclusion, the present study indicates that the majority of genetic effects on cardiometabolic risk factors remain relatively constant over middle age, with the noted exception of some specific genetic effects on ApoB and TG, which play a less prominent role in old versus young age.

## Data Availability

All data produced in the present study are available upon reasonable request to the authors

## Acknowledgments

The present study has been conducted using the UK Biobank Resource (Application Number 56340) that is available to researchers. We acknowledge supports from the Copenhagen General Population Study and the Estonian Biobank. This study was supported by the China Scholarship Council (CSC; no. 202106240064; to L Ao).

## CRediT authorship contribution statement

**Linjun Ao:** Conceptualization, Methodology, Formal analysis, Visualization, Writing – Original Draft, Writing-Review & Editing. **Raymond Noordam:** Conceptualization, Data Curation, Methodology, Writing-Review & Editing, Supervision. **Diana van Heemst & Ko Willems van Dijk:** Conceptualization, Writing-Review & Editing, Supervision. **Jiao Luo & Ruth Frikke-Schmidt & Maris Teder-Laving & Reedik Mägi:** Data Curation, Writing-Review & Editing. **Estonian Biobank Research Team**: data collection, genotyping, quality control, imputation. All authors have reviewed and approved the final version of the manuscript.

## Data and code availability

Data and code used in the presents study will be made available upon request in adherence with transparency conventions in medical research and through reasonable requests to the corresponding author.

